# Evaluating the Sensitivity of Event-Based Surveillance and Timeliness of Event and Indicator-Based Surveillance Systems in Kenya Using the 7-1-7 Metrics: A Multi-County Analysis, 2021–2025

**DOI:** 10.64898/2026.08.06.26359848

**Authors:** Stella Mamuti, Philip Ngere, Dalmas Omia, Mercy Mulaku, Eric Osoro, M Kariuki Njenga, Peninnah Munyua, Isaac Ngere

## Abstract

**Background:** Timely detection and response to public health threats are essential to mitigating outbreaks. Kenya adopted a phased rollout of Event-Based Surveillance (EBS) in May 2021 to complement Indicator-Based Surveillance (IBS). We evaluated the sensitivity of EBS and the timeliness of both EBS and IBS systems for confirmed outbreaks across four Kenyan counties.

**Methods:** We conducted a retrospective analysis of outbreak data from May 2021 to May 2025 in four counties in Kenya. We administered a structured questionnaire with the following variables: confirmed disease outbreak, dates of emergence, detection, notification, and initiation of each early response action, and the surveillance system that detected it. We also included bottlenecks and enablers for timely detection, notification, and response. The EBS sensitivity was summarized as a proportion, with the numerator being the outbreaks detected through EBS and total outbreaks as the denominator. Timeliness was measured using the 7-1-7 target, a global benchmark for measuring the timeliness of outbreak detection and response, where outbreaks should be detected within 7 days of disease emergence, notified within 1 day of detection, and initial response actions completed within 7 days of notification. The bottlenecks and enablers were summarized in a table.

**Results:** We recorded a total of 28 confirmed outbreaks across the four years, of which 16 (57.1%, 95% C.I.: 34–72%) were detected through EBS. A total of 15 (53.6%) met the full 7-1-7 metrics. Among EBS-detected outbreaks: 100.0% were detected within 7 days of emergence, 81.3% were notified within 1 day after detection, and 68.8% had initial response activities completed within 7 days after notification. For IBS, 83.3% were detected within 7 days, 75.0% notified within 1 day, and 75.0% had initial response activities completed within 7 days. Five of the seven (71.4%) Anthrax outbreaks met all the metric targets. Detection and notification were enabled by community systems (trained health promoters, linkages, and awareness), information systems (hotlines, dashboards, and digital reporting), and response systems (sample referral, and rapid response teams). Bottlenecks included limited reach, low suspicion, stigma, reporting downtimes, delayed escalation, and constrained response capacity due to funding, transport, personnel, and laboratory delays.

**Conclusion:** The results demonstrate that EBS contributes significantly to outbreak detection and notification, while IBS contributes more to response. Therefore, there is a need to leverage EBS’s strength in early detection and notification, and IBS’s better timeliness in response for a more timely system.

## Introduction

Timely detection and response to disease outbreaks are essential for effective public health surveillance. To strengthen early warning and response capacities, the revised International Health Regulations (IHR 2005) prompted WHO Member States to adopt both Indicator-Based Surveillance (IBS) and Event-Based Surveillance (EBS) as core components of the Integrated Disease Surveillance and Response (IDSR) strategy^1^. IBS focuses on routine case reporting using standard case definitions, while EBS emphasizes the immediate detection and reporting of unstructured “signals” that may indicate a public health threat^2^. The design of EBS in Kenya aligns with the Africa CDC framework and WHO Joint External Evaluation (JEE) indicators, both of which underscore the need for strong early warning and response (EWAR) capabilities across all levels of the health system^3,4^.

Although IBS and EBS are distinct, they are complementary. The structured disease-specific surveillance by IBS is well-suited for monitoring endemic trends and confirming outbreaks through laboratory data. However, it may fail to capture events in communities with limited access to health services or in communities where the proportion of the population that seeks formal healthcare is low. By contrast, EBS uses non-traditional sources, including community reports, media scanning, and hotline alerts, to detect early signs of potential outbreaks^5^. This approach allows for broader event capture and faster escalation. Since events precede most disease outbreaks, EBS is predictive and provides a lead time for putting measures in place to prevent or reduce adverse outcomes of the outbreak.

In Kenya, EBS was piloted in 2019 by the Ministry of Health, in collaboration with the US Centers for Disease Control and Prevention and other partners. Community Event-Based Surveillance (CEBS) was piloted in Siaya and Nakuru counties from September 2019 to May 2020, while Health Facility-level Event-Based Surveillance (HEBS) was piloted in three counties (Mombasa, Nakuru, and Meru). A phased national rollout began in 2021 across four counties: Meru, Mombasa, Nakuru, and Siaya. The counties were selected for their epidemiological diversity and readiness. In May, Busia County was added due to a high cross-border disease transmission risk, thus a strategic cross-border surveillance point^6^. Kenya’s EBS system was designed to operate within the existing IDSR framework, ensuring alignment with national surveillance priorities and global health security goals. The system follows a standardized cascade: signals are triaged, verified, subjected to risk assessment, and escalated for response if confirmed to be true events^2^. This structure enables efficient prioritization of threats while reducing false alarms. Early evidence suggests that EBS contributed to the prompt detection of specific outbreaks, such as clusters of severe acute respiratory illness^7^. However, there is limited data on other core performance metrics, particularly sensitivity and timeliness, essential to evaluating the systems’ effectiveness.

As Kenya scales up EBS rollout nationally, a comprehensive assessment is needed to determine whether this investment has yielded the intended improvements in complementing IBS in outbreak detection and response. The “7-1-7” benchmark proposed by Resolve to Save Lives and adopted by WHO offers a standard to assess timeliness^8,9^. The metric assesses whether detection occurs within 7 days of the outbreak emergence, notification is made within 1 day of detection, and early response actions are completed within 7 days of notification. The benchmark has been used in several settings to evaluate outbreak performance^10^. Such evidence is critical to inform strategic decisions on resource allocation and surveillance system strengthening at both national and sub-national levels. Here, we aimed to determine the sensitivity of EBS and to assess the timeliness of detection, notification, and response for outbreaks across both the EBS and IBS surveillance systems, drawing data from four Kenyan counties, which were among the first counties to roll out EBS: Busia, Meru, Nakuru, and Siaya.

## Methods

### Study Design and Setting

We conducted a retrospective review of confirmed outbreaks and the corresponding EBS signals from May 2021 to May 2025.

### Study Site

We conducted the outbreak evaluation in four counties, which were among the first counties to roll out EBS. The counties included: Siaya, Meru, and Nakuru, in which the EBS rollout started in May 2021, and Busia, which started the rollout in May 2022 (Figure 1). The study period was from the time the EBS rollout started in the county to May 2025.

**Figure 1.**
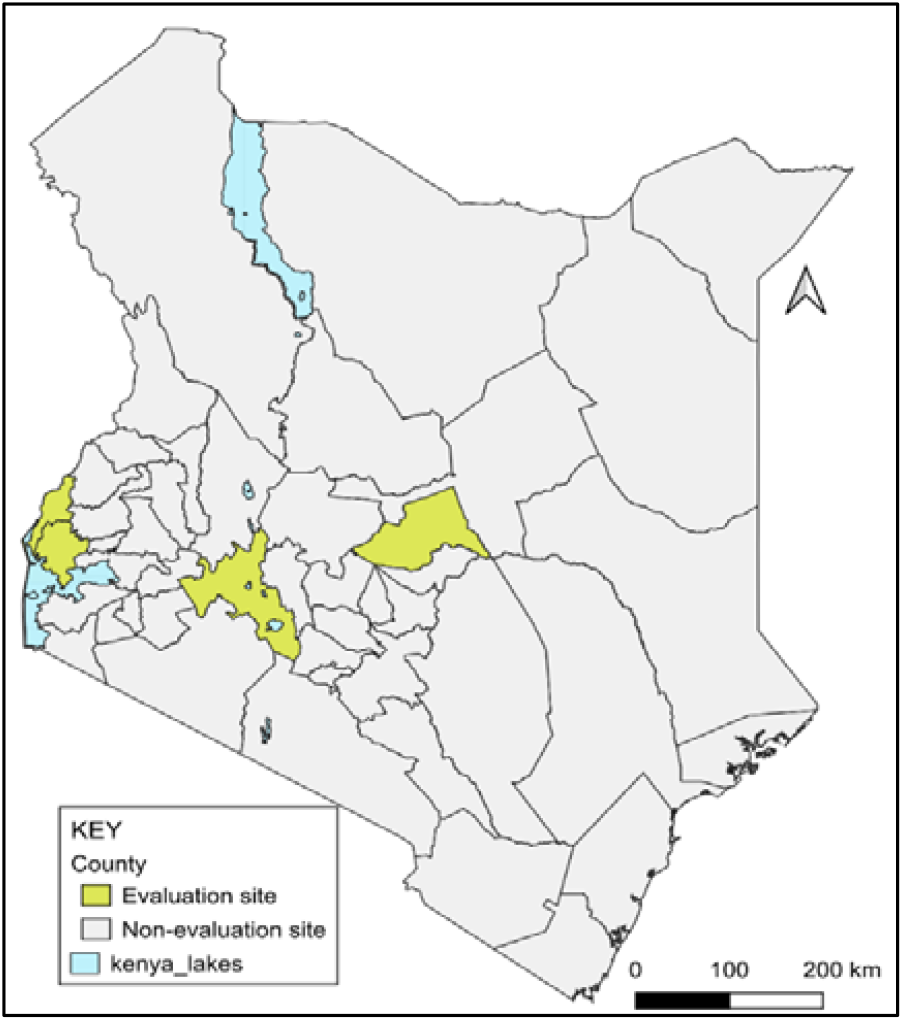
Study sites for evaluation of the sensitivity and timeliness of Event and Indicator-based surveillance systems

### Operational definitions

#### EBS outbreak detection

Confirmed outbreaks that had a corresponding signal in the EBS system. The outbreak was considered detected by EBS if the date of onset of symptoms, place of residence, and symptoms of the index case in the line list were associated with the type of signal reported, signal description, place/source of signal, and confirmation of the signal as true by the Community Health Assistant in the EBS system.

#### IBS outbreak detection

Confirmed outbreaks without a corresponding EBS signal at the time of detection and detected through formal health systems such as health facilities and laboratories.

#### Signal

Raw data and/or information reported through the EBS system representing a potential acute health risk, such as an outbreak. There are standard signals currently being used in Kenya, divided into Community EBS and Hospital EBS signals reported through a system called m-Dharura ^11^. There are other reporting systems, such as Hotline and phone calls, that include all events of Public Health concern, like disease outbreaks.

#### Outbreak

An occurrence of disease greater than would normally be expected in a defined area or among a specific group of people over a particular period. An outbreak of the same disease in a geographic area was considered new after a duration equivalent to at least two maximum incubation periods following the resolution of the last case in the previous outbreak.

#### Timeliness

The speed between steps in the outbreak containment process, that is, detection, notification, and response for the identified outbreaks.

#### 7-1-7 matrix

This is a global benchmark for measuring the timeliness of outbreak detection and response. The outbreak is detected within 7 days from the date of disease emergence, notified within 1 day of detection, and initial response actions completed within 7 days of notification^12^.

Where:

- **Date of emergence**
- For endemic diseases: a date on which a predetermined increase in case incidence over baseline rates occurred.
- For non-endemic diseases: the date on which the index case or first epidemiologically linked case first experienced symptoms.
- **“7” (Detection):** Identification of an outbreak within 7 days of symptom onset or event occurrence. The time between the date of emergence and the corresponding signal reporting in EBS, or the date the index case was seen at the facility for IBS.
- **“1” (Notification):** Notification of the outbreak to the relevant public health authorities within 1 day of detection.
- EBS: time between the date of signal reporting and notification to the Public Health authority (Sub-County Disease Surveillance Coordinator)
- IBS: time between the index case first seen at the facility and notification to the Public Health authority (Sub-County Disease Surveillance Coordinator)
- **“7” (Response):** Completion of initial public health response actions within 7 days of notification. The actions include: initiation of investigation or deployment of response team, epidemiological analysis and initial risk assessment, laboratory confirmation of the outbreak etiology, initiation of appropriate case management and infection prevention and control measures in health facilities, initiation of appropriate public health countermeasures in affected communities, initiation of appropriate risk communication and community engagement activities, and establishment of a coordination mechanism.

### Data collection

We began by enumerating outbreaks that had occurred from May 2021 to May 2025 in the study counties. Any outbreak investigations initiated at the county or national level, regardless of the disease, were listed. The County-level IDSR data for 2021-2025 for the listed disease outbreaks were re-analyzed to determine if outbreak definitions were met, as defined in the Kenya IDSR 3rd edition guidelines^2^.

Inclusion and exclusion criteria

All outbreaks that met the Kenya IDSR guidelines definition of an outbreak by disease were included in the study.

Outbreaks with incomplete key surveillance data, including missing dates for detection, notification, or initial response action, making 7-1-7 assessment impossible, were excluded.

We used a structured questionnaire to collect data about each of the identified outbreaks. We administered the questionnaire to the County and Sub-County Disease Surveillance Coordinators, who are government officers responsible for detection and response to disease outbreaks. The variables collected included confirmed disease outbreak, dates of emergence, detection, notification, early response actions, and the surveillance system that detected it. We also included bottlenecks and enablers for timely detection, notification, and response. Once the date of emergence of the outbreak was identified, reported signals from the same sub-county were cross-checked to identify if a signal related to the outbreak was reported within one month before the reported date of emergence. The outbreak was detected by EBS if the date of onset of symptoms, place of residence, and symptoms of the index case in the line list were associated with the type of signal reported, signal description, place/source of signal, and confirmation of the signal as a true event by the Community Health Assistant in the EBS system.

### Data Analysis

We summarized the data using descriptive statistics. We used proportion for EBS sensitivity where the number of confirmed outbreaks detected by EBS was the numerator, and the total confirmed outbreaks was the denominator. Calculated as: *(Number of outbreaks detected by EBS / Total number of outbreaks)* X *100*

We determined the timeliness for both the IBS and EBS detection, notification, and response, and mapped them to the 7-1-7 metrics. Timeliness was considered achieved if all the 7-1-7 targets were met. The qualitative data were summarized into enablers and bottlenecks for detection and notification, and response.

### Ethical considerations

We received study approval from the Kenya Medical and Research Institute (KEMRI) Scientific and Ethics Review Unit (SERU) No. 5094. A letter was written to the Ministry of Health, Division of Disease Surveillance and Response, to request access to the data. Permission was sought from the County administration before proceeding with the interviews.

### Results Sensitivity

Across the four study counties, 28 outbreaks were confirmed between May 2021 and May 2025. Anthrax and Cholera each accounted for 25.0% (7/28) of all outbreaks. The Event-Based Surveillance detected 16 of the 28 confirmed outbreaks, translating to a sensitivity of 57.1% (95% C.I.: 34–72%). The EBS sensitivity varied by disease, highest for anthrax (5/7, 71.4%) and lowest for Monkeypox (2/4, 25.0%) (Table 1**Error! Reference source not found**.).

**Table 1:**
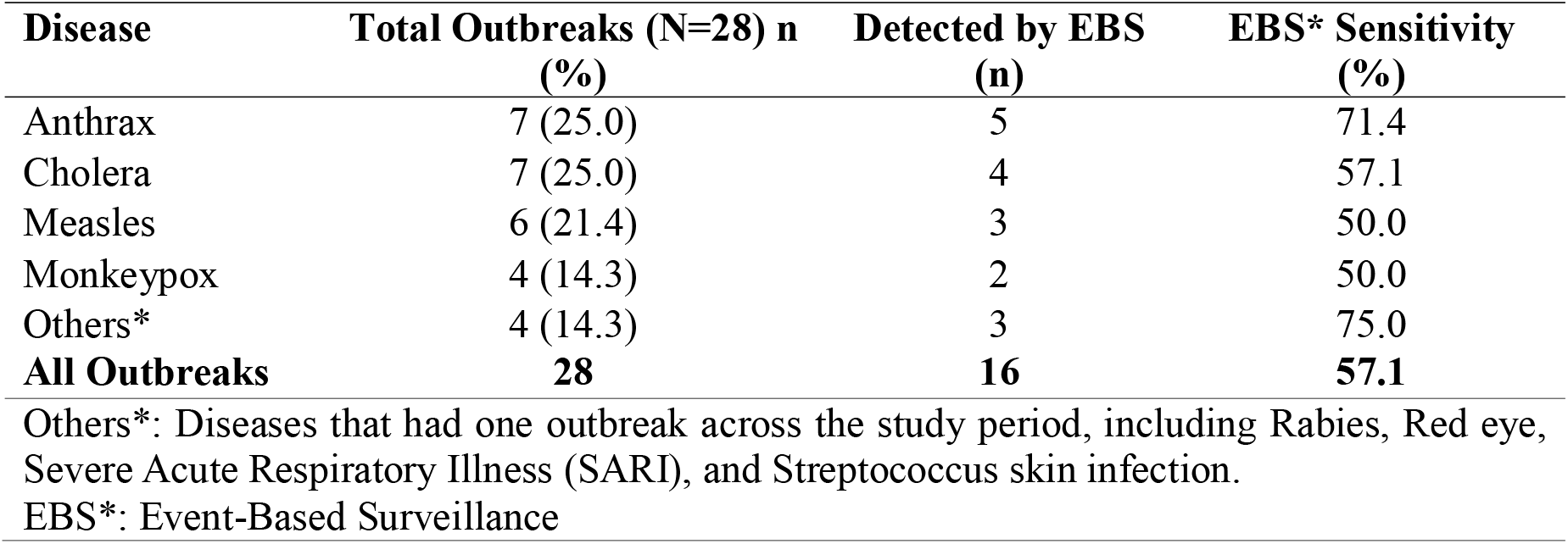
Sensitivity of Event-Based Surveillance System in outbreak detection in selected counties, Kenya, May 2021–May 2025.

| Disease | Total Outbreaks (N=28) n (%) | Detected by EBS (n) | EBS* Sensitivity (%) |
| --- | --- | --- | --- |
| Anthrax | 7 (25.0) | 5 | 71.4 |
| Cholera | 7 (25.0) | 4 | 57.1 |
| Measles | 6 (21.4) | 3 | 50.0 |
| Monkeypox | 4 (14.3) | 2 | 50.0 |
| Others* | 4 (14.3) | 3 | 75.0 |
| **All Outbreaks** | **28** | **16** | **57.1** |

Others\*: Diseases that had one outbreak across the study period, including Rabies, Red eye, Severe Acute Respiratory Illness (SARI), and Streptococcus skin infection. EBS\*: Event-Based Surveillance

**Table 2:** Timeliness of Detection, Notification, and Response by Surveillance System, Kenya, 2021–2025.

| 7-1-7 Metric | *EBS (N=16) | *IBS (N=12) | All outbreaks (N=28) |
| --- | --- | --- | --- |

|  | n (%) | n (%) | n (%) |
| --- | --- | --- | --- |
| Detection $\leq 7$ days | 16(100.0) | 10(83.3) | 26(92.9) |
| Notification $\leq 1$ day | 13(81.3) | 9(75.0) | 22(78.9) |
| Response $\leq 7$ days | 11(68.8) | 9(75.0) | 20(71.4) |
| **Met Full 7-1-7** | 9(56.3) | 6(50.0) | 15(53.5) |

\*EBS: Event-Based Surveillance
\*IBS: Indicator-Based Surveillance

### Timeliness by 7-1-7 Metric

The proportion of all the recorded outbreaks that met the 7-1-7 target was 53.5% (15/28). Of the EBS-detected outbreaks, 9 of 16 (56.3%) met the 7-1-7 target, while 6 of 12 (50%) of the IBS-detected outbreaks met the target. Among EBS-detected outbreaks: 100% were detected within 7 days, 81.3% were notified within 1 day after detection, and 68.8% had initial response activities completed within 7 days after notification. For IBS: 83.3% were detected within 7 days, 75% notified within 1 day, and 75% had initial response activities completed within 7 days were 75% (**Error! Reference source not found.Error! Not a valid bookmark self-reference**.).

### Timeliness by disease type

Two (50%) of the Monkeypox outbreaks detected through IBS did not meet the 7-day target for detection. Two measles and one each of Monkeypox, Cholera, Red Eye, and Severe Acute Respiratory Illness (SARI) outbreaks were notified beyond the 1-day target. Three Measles (50%), three Cholera (43%), and two Anthrax (29%) had the initiation of the initial response activities completed after the 7-day target (Figure 2: Timeliness of Event-Based and Indicator-Based Surveillance Systems by Disease Outbreak in Selected Counties, Kenya, 2021-2025. The y-axis represents the number of days).

**Figure 2.**
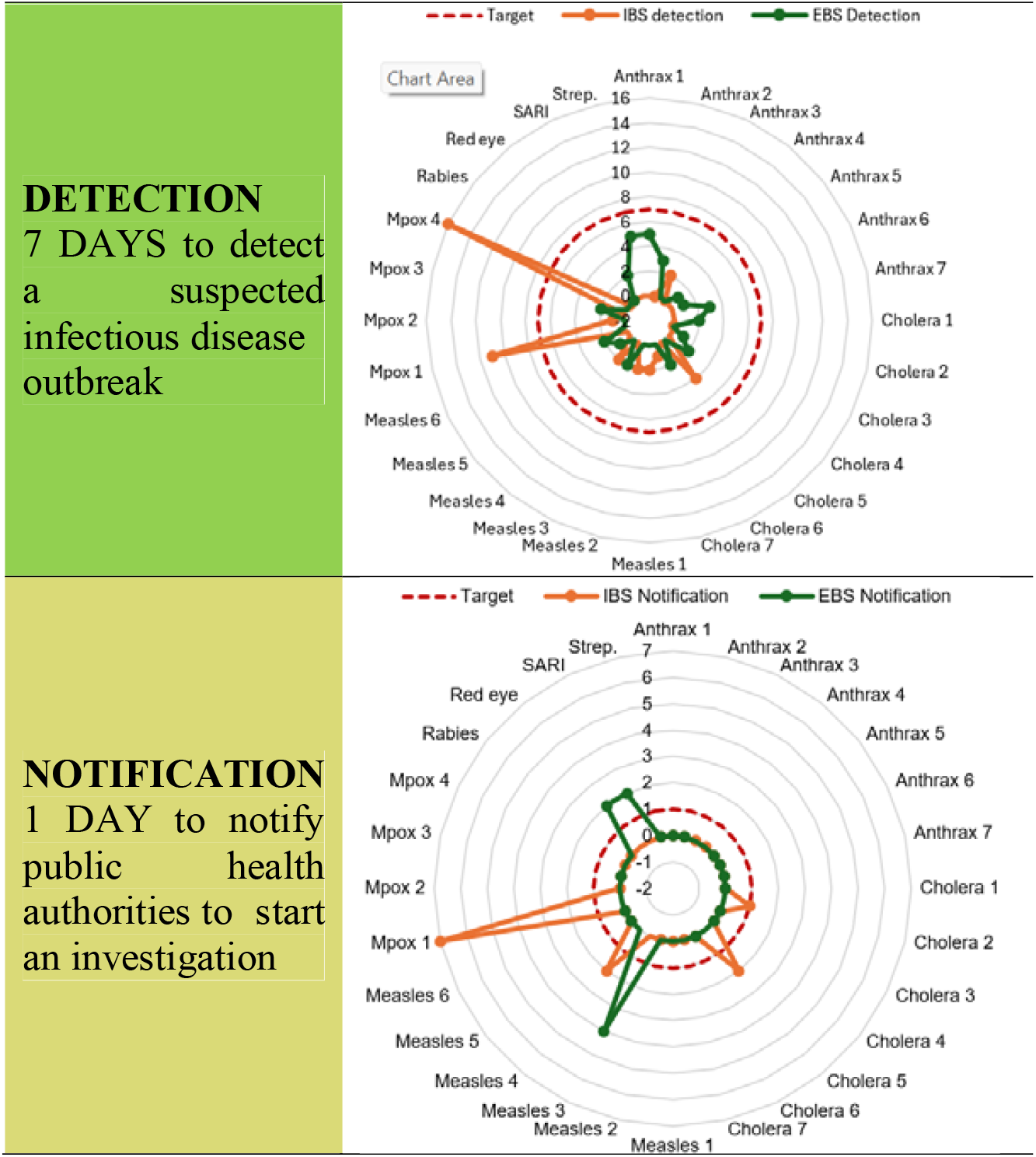
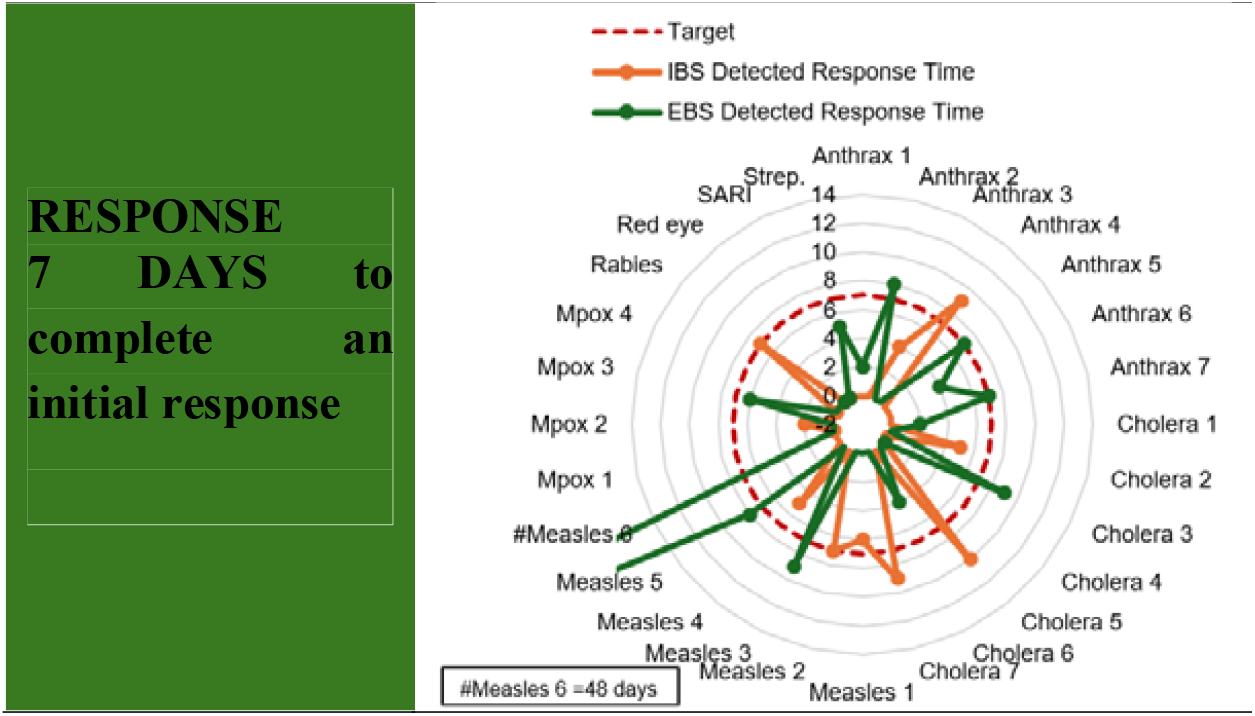
Timeliness of Event-Based and Indicator-Based Surveillance Systems by Disease Outbreak in Selected Counties, Kenya, 2021-2025. The y-axis represents the number of days

### Enablers and Bottlenecks to Timely Detection, Notification and Response

Several enablers and bottlenecks to timely detection, notification, and response were identified. Detection and notification were enabled by community systems (trained health promoters, linkages, awareness), information systems (hotlines, dashboards, digital reporting), and response systems (sample referral, rapid teams). Bottlenecks included limited reach, low suspicion, stigma, reporting downtimes, delayed escalation, and constrained response capacity due to funding, transport, personnel, and laboratory delays (*Table 3*).

**Table 3:** Facilitators and barriers to timely Detection, Notification, and Response.

| Category | Enablers | Bottlenecks |
| --- | --- | --- |
| Detection and Notification | 1. Trained Community Health Promoters (CHPs) on Event-Based Surveillance protocols<br>2. Functional community linkage systems<br>3. Public awareness of disease symptoms<br>4. Port health screening (e.g., for Monkeypox)<br>5. Healthcare workers' knowledge of notifiable diseases<br>6. Timely feedback from the national Public Health Emergency Operation Center<br>7. Use of 719 hotline for self-reporting by patients | 1. Unlinked closed/migratory populations<br>2. Low index of suspicion among the community and healthcare workers<br>3. Stigma (e.g., Monkeypox associated with sex workers)<br>4. Weak cross-country surveillance linkage, limiting the timely detection of epidemiologically linked cases<br>5. Reporting system downtime - Delayed transmission of signals to the Community Health Assistant for verification<br>6. Community Promoters fear being blamed in the community for reporting certain diseases (e.g., anthrax cases related to consumption of condemned carcasses) |
| Response | 1. Availability of trained rapid response teams at both national and sub-national levels<br>2. Availability of emergency supplies<br>3. Functional sample referral system<br>4. Community support for mobilization<br>5. WhatsApp groups and dashboards for coordination of response activities<br>6. Inter-departmental coordination | 1. Competing priorities delaying risk assessments<br>2. Delayed emergency fund disbursement<br>3. Limited transport for field deployment<br>4. Shortage of skilled personnel (e.g., for sample collection)<br>5. Long turnaround times for laboratory results |

## Discussion

Our findings show that EBS had a moderately high sensitivity and high timeliness in outbreak detection. The ability of EBS to detect outbreaks early, particularly those of sudden onsets such as Cholera and cross-sectoral outbreaks such as anthrax, demonstrates its value in resource-constrained environments.

### Sensitivity

Overall, EBS demonstrated moderate sensitivity, detecting more than 50% of the outbreaks. This finding aligns with previous studies, such as evaluations in Cambodia and Ethiopia, where EBS detected between 45–60% of confirmed outbreaks depending on the strength of community-based surveillance systems and the functionality of reporting mechanisms^13^. The EBS sensitivity was highest for outbreaks that had high community-level suspicion index, particularly those with visible symptoms or public concern, such as anthrax. The high suspicion index might have been enhanced by the integration of community-based actors such as Community Health Promoters (CHPs), animal health workers, and village elders in Kenya’s EBS model trained on notifiable diseases ^14,13,15,16^.

The findings reveal that while EBS holds promise as an early detection system, its sensitivity varies across specific disease outbreaks. For example, low community awareness and stigma surrounding certain diseases, e.g., Monkeypox, can suppress reporting, as seen in previous studies from West Africa and Southeast Asia^17,18^. At the onset of the Monkeypox outbreak in Kenya, the low community awareness about the signs and symptoms of Monkeypox, being a reemerging disease, made it hard for the disease to be detected at the community level through EBS. After the community became more aware of Monkeypox, stigma associated with the presenting symptoms and mode of transmission hindered detection by EBS in the study counties. Continuous support, especially in sensitization of CHPs on emerging and re-emerging diseases, and feedback mechanisms, can enhance EBS sensitivity. In the absence of such support, signals may be overlooked, dismissed, or not escalated, especially when the clinical presentation is nonspecific or when reporters lack confidence in system responsiveness.

### Timeliness

More than half of all outbreaks met the full 7-1-7 criteria, with variation across the EBS and IBS systems and outbreak type. This performance is higher than the performance in similar contexts, where full metric compliance was found to be 27%, with variations across countries and specific disease outbreaks^19^. This finding suggests that even though the 7-1-7 target is feasible, full compliance is affected by several factors, such as geographical regions, type of outbreak, surveillance system, and availability of resources. The 7-1-7 metric can therefore be used not only as an evaluation tool but as a quality improvement tool for measuring the resilience of a surveillance system across different locations.

Event-Based Surveillance showed a higher proportion in meeting the full 7-1-7 metric. The faster detection and notification associated with EBS may be attributed to earlier community-level recognition and quicker local escalation, especially when empowered by functional alert management and triage teams. These findings reinforce the added value of EBS not only in sensitivity but also in enabling faster public health event detection and notification, particularly in detecting syndromes or unusual events that may not immediately present to healthcare facilities ^20,21^. However, the proportion of EBS-detected outbreaks that had the initial response activities initiated within 7 days was lower compared to those detected by IBS. This is likely because IBS is based on predefined thresholds for responding to notifiable diseases, allowing for faster formal response initiation^19,23^. To address this gap, there is a need for stronger interoperability between EBS and IBS systems. Specifically, once an EBS signal is verified as a true event or outbreak, it should automatically reflect in the IBS reporting structure. By combining the early detection capabilities of EBS with the rapid activation mechanisms of IBS, the national surveillance system would become more agile and responsive.

Anthrax and cholera outbreaks had a median detection of 2 days, and most anthrax outbreaks met the full 7-1-7 criteria. This performance may be attributed to a combination of factors, including the acute and easily recognizable clinical presentations of both diseases. Observable signs such as sudden skin lesions and swelling in anthrax, and profuse watery diarrhea in cholera are more likely to prompt suspicion and immediate action by both community members and healthcare workers. In addition, the EBS reporting system includes signals on sudden livestock deaths or acute watery diarrhea, which are likely to speed up detection and reporting ^20^. The early detection and response for the two diseases may also reflect the historical prioritization of these diseases. Under Kenya’s IDSR strategy, both anthrax and cholera are classified as notifiable priority conditions requiring immediate reporting ^2,22^. Regular training of healthcare workers on case definitions, outbreak thresholds, and response protocols for these conditions under IDSR has likely reinforced timely reporting behaviors ^23^. Globally, similar patterns have been observed. In Uganda and Tanzania, for instance, diseases like anthrax and cholera are more rapidly reported and responded to than less familiar or stigmatized conditions, due to better clinical recognition and greater public health infrastructure preparedness ^24,25^.

Monkeypox outbreaks had a slightly longer detection period, suggesting delayed identification compared to other disease outbreaks. Stigma associated with Monkeypox due to its visible lesions, associations with sexual transmission, and recent links to marginalized groups were the likely causes of late detection. This finding is similar to the findings during the 2022–2023 global outbreak, where Monkeypox was heavily stigmatized in both high- and low-resource settings, especially when early cases were concentrated among men who have sex with men ^26,27^. The stigmatization led to underreporting, misinformation, and reduced care-seeking, which in turn hindered early detection by both EBS and IBS mechanisms^27^.

Measles outbreaks experienced substantial delays in initiation of response activities. The delayed response might be due to weaknesses in emergency coordination and possibly resource constraints for initiating mass immunization catch-up campaigns. This delay is consistent with trends reported across many low- and middle-income countries (LMICs), where measles outbreak response was often delayed by the logistical complexity of vaccination campaigns^28^.

### Study limitations

A key limitation of this retrospective study is the potential for recall and documentation bias, especially regarding the accuracy of dates of initiation of early response actions. However, this bias was likely minimized through reliance on routinely maintained digital reporting systems used by both EBS and IBS platforms.

## Conclusion

The results demonstrate that EBS contributes significantly to enhancing the timeliness of outbreak detection and notification, but its response was lower compared to IBS. Therefore, there is a need to continue leveraging EBS’s strength in early detection and notification, and IBS’s better timeliness in response, for more coordinated and timely responses across both informal and formal data streams, limiting the spread of infectious diseases. In addition, there is a need for targeted interventions to overcome the identified bottlenecks in order to improve timeliness.

## Data Availability

De-identified data that support the findings of this study will be made available upon reasonable request from the corresponding author after publication. Data will be shared in accordance with applicable ethical and data governance requirements.

## Authors’ Contributions

SM, IN, PN, and PM conceived the evaluation and contributed to the evaluation designs; SM, IN, and NK supported acquiring, managing, and interpreting the data, and accessed and verified the data; SM, IN, and PN supported the analysis; SM prepared the manuscript, and all authors contributed to the revision of the manuscript. All authors were responsible for submitting the manuscript for publication.

## Acknowledgments

We acknowledge the Kenya National Public Health Institute (KNPHI) for technical leadership, coordination, and support throughout the implementation of this work. We also acknowledge the County Departments of Health and surveillance teams in Busia, Meru, Nakuru, and Siaya counties for their collaboration, facilitation, and provision of data and insights that informed this study. We are grateful to all surveillance officers, health facility staff, community health promoters, and emergency operations personnel who contributed to detection, reporting, verification, and response activities in the study sites.

